# Treatment provision for adults with ADHD during the COVID-19 pandemic: An exploratory study on patient and therapist experience with on-site sessions using face masks vs. telepsychiatric sessions

**DOI:** 10.1101/2020.12.11.20242511

**Authors:** Helen Wyler, Michael Liebrenz, Vladeta Ajdacic-Gross, Erich Seifritz, Susan Young, Pascal Burger, Anna Buadze

## Abstract

**Background:** Maintaining the therapeutic care of psychiatric patients during the first wave of the COVID-19 pandemic in Switzerland required changes to the way in which sessions were conducted, such as telepsychiatric interventions or the use of face masks during on-site sessions. While little is known about how face masks affect the therapeutic experience of patients and therapists, the effectiveness of telepsychiatry is well documented for several psychiatric disorders. However, research on the benefits of telepsychiatry in adult patients with attention-deficit/hyperactivity disorder (ADHD) remains scarce. This seems problematic since the symptoms typically associated with ADHD, such as attention problems and distractibility, may lessen the utility of telepsychiatry for this particular group. The present study’s aim was to explore how adult patients with ADHD and their therapists experienced therapy sessions during the COVID-19 pandemic in three different settings: face-to-face with the therapist wearing a face mask, via telephone, or via videoconferencing.

**Methods:** In this exploratory, quantitatively driven mixed-method study (quantitative questionnaire data and qualitative data from open-ended responses), we assessed patients’ evaluation of the session, their treatment satisfaction, and patients’ and therapists’ ratings of therapeutic alliance. We also collected qualitative comments on both sides’ experience of the session. Overall, 97 therapist and 66 patient questionnaires were completed. Results are reported for the *N* = 60 cases for which data from both parties were available. Sequential multiple regressions adjusted for therapist and number of sessions were used for the main quantitative analyses.

**Results:** Telepsychiatric sessions were rated as significantly less deep than face-to-face sessions, an effect that was mainly driven by lower ratings in the videoconferencing group and, as suggested by further analyses, may decline over time. No other statistically significant differences were observed. Elements that were mentioned as facilitating or complicating a session differed markedly between patients and therapists.

**Conclusions:** Both settings, on-site with the therapist wearing a face mask and telepsychiatric, seem to be valid options to continue treatment of adults with ADHD during a situation such as the COVID-19 pandemic. Aspects such as patient preference, session content, and therapeutic methods may be useful to identify the most suitable modality.

---

The first Swiss COVID-19 case was confirmed on the 24^th^ of February 2020 [3]. On the 16^th^ of March, the Swiss Federal Council declared an ‘extraordinary situation’ and introduced stringent regulations to slow down the spread of the virus [COVID-19 Ordinance 2; 4] during the so-called first wave; citizens were advised to stay home whenever possible and gatherings of more than five individuals were prohibited shortly thereafter. The impact of the COVID-19 pandemic was not confined to people’s working and social lives, however. First and foremost, it affected their access to health care, including psychiatric care. Following the declaration of the ‘extraordinary situation’, medical treatment providers, including psychiatrists and psychotherapists, were obliged by the government not to offer treatment that was not considered to be medically urgent in order to decelerate the spread of the virus [5]. At the same time, psychiatrists and psychotherapists had a duty to maintain psychiatric and psychotherapeutic care of the Swiss population by delivering indicated treatment that could not be delayed [see e.g. 6].

To cope with this dilemma, both the society of Swiss psychiatrists – the *Foederatio Medicorum Psychiatricorum et Psychotherapeuticorum* [7] – and the Confederation of Swiss Psychologists [8] recommended that treatment via telephone or videoconferencing tools should be implemented and chosen over face-to-face sessions. In other words, psychiatrists and psychotherapists were advised to resort to telepsychiatric treatment whenever possible, even though this form of treatment provision was only partially reimbursed by health insurers [9]. If on-site treatment was necessary and unavoidable, significant changes to the course of a session had to be made. Based on the recommendations of the Federal Office of Public Health and the FMPP [10], the internal regulations of the outpatient clinic in which the present study was conducted included: (1) following hygiene rules before the session (supervised hand disinfection and temperature measurement including a check for symptoms of a possible COVID-19 infection at the entrance of the building), (2) observing distancing rules during the session, and (3) the therapist wearing a face mask during the session. Thus, continuing psychiatric or psychotherapeutic treatment during the COVID-19 pandemic required substantial changes to the way sessions were usually conducted, either by resorting to telepsychiatry or by making significant adjustments to on-site sessions.

## The use of face masks in face-to-face treatment

Thus, continuing psychiatric or psychotherapeutic treatment during the COVID-19 pandemic required substantial changes to the way sessions were usually conducted. In addition to telepsychiatry, on-site sessions are also conceivable, especially if face masks are used in addition to strict adherence to hygiene and distance rules. While questions relating to the potential effects wearing face masks might have on therapy sessions and the therapeutic relationship have been discussed in recent Letters to the Editor [11-14] and discussion papers [15], little is known about their actual impact. A randomised controlled trial from Hong Kong in 2011, which examined patients’ reaction to their general practitioner wearing a face mask during the consultation, found negative effects on patients’ perception of the general practitioner’s empathy, an effect that was more pronounced for more established general practitioner-patient relationships [16]. We are not aware of a study that has investigated psychiatric outpatients’ experience of the session or the therapeutic relationship when the therapist is wearing a face mask.

## Telepsychiatry versus face-to-face therapy

A main benefit of *telepsychiatry* – in this context defined as synchronous therapeutic interventions that are not delivered on-site – is often seen in its ability to provide access to care for patients who, for instance, live in rural or remote areas [see e.g. 17, 18]. However, the COVID-19 pandemic highlighted the importance of telepsychiatry in upholding provision of treatment independently of physical location in general.

Over the past decades, the effectiveness of telepsychiatry has been examined in numerous studies. With respect to treatment outcome, empirical evidence suggests that telepsychiatry is on a par with face-to-face treatment for various psychiatric disorders [19], including anxiety disorder [20, 21], depression [21], and post-traumatic stress disorder [22-24]. Also, patients seem to be as satisfied with tele-mental health interventions as they are with in-person treatment [25, see also e.g. 26]. Some therapists, however, have expressed concerns that the lack of physical interaction in a telepsychiatric setting could hinder the development of a healthy *therapeutic alliance* (TA) [see e.g. 27], which is often conceptualised as (1) an emotional bond between patient and therapist and (2) collaboration and consensus between the two parties on the goals and tasks of the therapy [28]. A thorough understanding of the impact of treatment modality on TA is essential given that TA has been found to be one of the strongest predictors of treatment outcome in face-to-face psychotherapy [29, 30]; despite this, the specific relevance of TA and its individual sub-components in telepsychiatric settings still warrants further investigation [31, 32]. Reviews of the literature suggest, however, that concerns about TA in telepsychiatry may be unfounded. Patients’ ratings of TA are fairly comparable for in-person and telepsychiatric interventions, for both video therapy [33] and therapy delivered via telephone [34], although results from the latter review may have been influenced by the fact that most of the studies included used non-randomised opportunity samples. Interestingly, SG Simpson and CL Reid [33] reported that therapists did not rate TA quite as highly as their patients, especially in the early stages of treatment. Across the different treatment modalities, some therapists seemed – at least initially – slightly more sceptical, whereas patients appeared to perceive TA similarly across the different treatment modalities.

## Attention-deficit/hyperactivity disorder (ADHD)

ADHD is a highly inheritable [35] neuropsychiatric disorder [36] characterized by its core symptoms of inattention, impulsivity and hyperactivity and further accessory symptoms such as emotional over-reaction and affective lability [37]. The view of ADHD as a disease of youth that is outgrown in later life [see e.g. 37] has had to be revised over the course of the last few decades. ADHD may persist either in a form that still meets the full criteria for a diagnosis of ADHD or in the form of clinically significant impairment associated with residual symptoms [38-41], as symptoms of hyperactivity and impulsivity generally tend to decrease with age [42]. The prevalence rate for ADHD up to the age of 18 is estimated at 5.3% [36] and for adult ADHD at 2.5% [43].

While ADHD is still highly prevalent in adulthood, it is also a severely underdiagnosed condition [44]. As recommended by the NICE guidelines, depending on the individual, a multimodal treatment, often including a combination of drug treatment and non-pharmacological treatment, should be offered. If left untreated, adult ADHD can be accompanied by severe functional impairments and limitations in multiple areas of life and is further associated with heightened risk of multiple mental health and social difficulties as well as premature mortality [45-49]. Some adults with ADHD succeed in developing compensation strategies to mitigate the impact of their symptoms on their work and social life [50]. However, these strategies may no longer be sufficient in situations of exceptional stress, such as the COVID-19 pandemic, which severely disrupted people’s everyday routine (e.g. introduction of home office or home schooling). It is therefore essential to ensure continuous availability of therapeutic support for patients with clinically relevant ADHD not only beyond childhood and adolescence, but also during such exceptional situations.

## Telepsychiatry for adults with ADHD

In their recent review of the literature, Spencer and colleagues [51] identified 11 relevant primary studies with sample sizes of at least 20 participants investigating the use of telemedicine in patients with ADHD. Overall, the results were promising; telepsychiatry was both well accepted and valued by patients and also associated with improved outcomes. However, only one of the studies also included an adult sample. Moreover, telemedicine was used either to augment standard care or for consultation or evaluation purposes, not as a (temporary) substitute for face-to-face treatment.

The question of the suitability of telepsychiatric treatment for (adult) patients with ADHD seems particularly pertinent because of typical impairments associated with this disorder. On the one hand, patients might find it more difficult to stay focused during an off-site session due to the abundance of potential distractors in their environment. They may also find it difficult to cope with the demands of a telepsychiatric session in terms of time management and self-organisation, as these skills usually tend to be impaired in ADHD. Moreover, the setting in itself, using a media device like a smart phone or computer, might lead to patients carrying out activities simultaneously to the psychotherapeutic session. Thus, we see tangible risks for increased distractibility with this form of psychotherapy in this specific group of patients. On the other hand, telepsychiatric interventions could counteract the negative effects of patients’ disorganisation, e.g. by saving travel time and thus alleviating experiences of stress due to difficulties in time management [52]. Furthermore, adults with ADHD may experience a range of financial problems [53], and telepsychiatry would allow those affected to save on costs associated with traveling to the therapy sessions. Hence, further research on how this specific group of patients experience telepsychiatric care is necessary.

## The Present Study

The aim of the present study was to explore how outpatients with ADHD and their therapists experienced therapy sessions under the adjustments necessary due to the COVID-19 pandemic. Each session took place either face-to-face with the therapist wearing a face mask, via videoconferencing, or via telephone; we subsume the latter two modalities under telepsychiatry. We intended to address two questions in particular. First, does telepsychiatric treatment offer a viable (temporary) alternative to face-to-face treatment in adult ADHD? As discussed, this question is important as several symptoms often associated with ADHD might result in telepsychiatry being particularly challenging for these patients. Second, how does on-site treatment with the therapist wearing a face mask compare to telepsychiatric treatment options? We are not aware of a comparative study investigating this question. Although explored in a specific group of patients, this examination of the impact of the therapist wearing a face mask will be of general interest.

Patients’ and therapists’ experience with the sessions were explored both quantitatively and qualitatively. We assessed patients’ experience and evaluation of the session, including their satisfaction with the therapy, and were also interested in patients’ and therapists’ ratings of TA. We further assessed whether patients felt the current sessions were comparable or inferior to the pre COVID-19 sessions and what setting they would prefer out of a number of different options. Finally, we asked patients and therapists about their experience of the session to obtain qualitative insights.

## Method

### Study Design

This exploratory, quantitatively driven mixed-method study (combining quantitative questionnaire data and qualitative data from responses to open-ended questions; [54-56]) with an independent measures design aimed to explore ADHD patients’ and therapists’ experience of a specific therapy session during the COVID-19 pandemic in one of the following three treatment modalities: face to face, videoconferencing, and telephone.

### Sample and Recruitment

Participants in this study were adults with a diagnosis of ADHD according to the 10^th^ revision of the International Classification of Diseases [57], who were receiving treatment (multi-modal therapy combining pharmacological therapy and psychotherapy) at a specialised outpatient clinic. Data were collected between the end of April and the end of June 2020^1^. The type of treatment modality during the pandemic was determined based on each patient’s needs and preferences. All patients with sufficient command of the German language and who attended a treatment session during the study period with one of the two participating therapists were offered the opportunity to participate in the research.

Overall, therapists completed 97 questionnaires. For 66 of these questionnaires a patient questionnaire was returned (response rate of 68%). Data from six patients had to be excluded, either because essential information was missing (*n* = 3), their data were collected at a point at which the wearing of a face mask was not mandatory for therapists (*n* =2), or because they did not have a diagnosis of ADHD (*n* = 1), yielding a final sample size of *N* = 60 (face to face: *n* = 29; videoconferencing: *n* = 11; telephone: *n* = 20). The two therapists involved in the study recruited similar numbers of patients (50 and 47) and had a similar response rate (66% and 70%). Thirty-one patients from Therapist A and 29 patients from Therapist B were included in the final sample. The two therapists conducted similar numbers of face-to-face and telepsychiatric sessions (*p* = .993), although, at a descriptive level, Therapist A conducted more sessions over the phone (12 vs. 8) whereas Therapist B ran more videoconferencing sessions (7 vs. 4).

### Measures

#### Session Evaluation Questionnaire

The *Session Evaluation Questionnaire* [SEQ; 58] is used to assess patients’ self-reported immediate session experience. It consists of 21 items scaled according to a semantic differential (e.g. from 1 *shallow* to 7 *deep*). The factor structure of the English original was replicated in the German version for the factors *session depth* and *session smoothness* (both in-session processes) and *post-session positivity* (post-session impact), but not for *arousal* [59]. Therefore, *arousal* was not included in the present study. Studies that investigated construct validity showed expectation-consistent associations with therapeutic alliance [59]. Higher scores indicate more depth, higher smoothness, and higher post-session positivity. Cronbach’s alpha values in the current study were satisfactory to good with .70 (*depth*), .77 (*smoothness*)^2^, and .87 (*post-session positivity*).

#### Client Satisfaction Questionnaire

The *Client Satisfaction Questionnaire* [CSQ; 60] is an eight-item self-report scale that assesses patient satisfaction with the mental health services indicated by means of 4-point Likert-type responses. The German version of the CSQ [61] has been reported to be reliable (Cronbach’s alpha = .90) and to possess good psychometric properties overall [62]. The instructions were slightly adapted for the current study, stating “when answering the following questions, please consider the therapy sessions as they took place today”. The mean score is reported with higher scores indicating higher satisfaction. Cronbach’s alpha was good (.84).

#### Working Alliance Inventory

The *Working Alliance Questionnaire* (WAI) is a common measure used to assess therapeutic alliance [33]. We applied the short revised version of the WAI for patients (WAI-SR-P) and therapists (WAI-SR-T) [63]. Both versions consist of 12 items; answers are provided on a 5-point Likert scale ranging from *seldom* (1) to *always* (5). The German version of the WAI-SR-P has good internal consistency, with Cronbach’s alpha ranging between .81 and .91 for the four subscales [64, 65]. For the German version of the WAI-SR-T, only limited data are available; for this study, the 12-item version recommended by T Munder [66] was used. The items of both WAI-SR questionnaires assess three subscales (*bond, tasks*, and *goals*). Mean scores are reported for each scale, with higher scores indicating a stronger working alliance. Cronbach’s alpha values for the WAI-SR-P were acceptable to good for *bond* (.76) and *tasks* (.80), but questionable for *goals* (.66). As deleting an item on the scale did not improve Cronbach’s Alpha, the scale was left unchanged. The corresponding values for the WAI-SR-T were acceptable at .72, .72, and .75, respectively.

#### Global Assessment of Functioning Score

The *Global Assessment of Functioning* (GAF) scale [67] was used to assess overall patient functioning from the therapist’s perspective. The GAF score ranges from 1 to 100, with 100 representing a high functioning individual that is not experiencing any symptoms.

#### Questions regarding the patient’s experience with the COVID-19 pandemic

A self-developed four-item questionnaire was used to assess patient experience with the COVID-19 pandemic. Patients were asked (1) whether they belonged to the high-risk group (yes/no) and to indicate the extent to which (2) their everyday life was affected by the COVID-19 pandemic, (3) they felt distressed because of the pandemic, and (4) they experienced fear because of the pandemic (all 5-point Likert-scales ranging from 1 *not at all* to 5 *very much*). The German questions and English translations can be seen from the Supplementary File 1 (Table A, questions C1-C4).

#### Specific closed and open-ended questions regarding the sessions (patient questionnaire)

Patients were asked whether they felt the current treatment modality was better, worse or comparable to the pre-COVID-19 therapy sessions, and how they would rank five treatment modality options (telephone; videoconferencing; face-to-face, in compliance with hygiene and distance rules; face-to-face with the therapist wearing a face mask; face-to-face with a plastic divider separating therapist and patient) in terms of preference if they had a free choice (see Supplementary File 1, Table A, questions F1 and F2). Moreover, patients were asked two open-ended questions: (1) “Was there anything you particularly liked about the way today’s session took place?”, and (2) “Was there anything you disliked about or felt uncomfortable with in the way today’s session took place?” (see Supplementary File 1, questions OP1 and OP2, for the original German wording).

#### Specific closed and open-ended questions regarding the sessions (therapist questionnaire)

Therapists were asked how many sessions patients had already had in the current modality and why that modality had been chosen (severity of the present condition, patient belonging to a risk group, patient preference, other). They were also asked how they evaluated the current session modality compared to the normal modality for this specific patient (open-ended question): “For this patient, how do you evaluate the way the therapy sessions are currently conducted compared to the way they normally take place?” (see Supplementary File 1, question OT1, for the original German wording).

### Procedure

Therapists informed eligible patients about the study at the end of a therapy session and provided them with the information sheet and their personal code. Patients were asked to complete the survey shortly after the session as the questions related to their experience of the therapy session; seventy percent of participants completed the survey on the same day. Data were collected through the online platform *Qualtrics* (Qualtrics, Provo, UT). Participants provided informed consent by ticking boxes to indicate that they had understood the aims of the study and their rights. The survey consisted of three questionnaires (SEQ, CSQ, and WAI-SR-P) and a number of questions on the patients’ experience with the session and the treatment modality, how the pandemic affected them in general, and demographics.

The therapists also received an information sheet and were asked to provide informed consent, as the completion of the WAI meant that they, too, were participants in this study. The therapists’ *Qualtrics* survey consisted of the WAI-SR-T, questions on their perception of the specific session, general questions regarding the session (reasons for modality chosen, number of sessions in the current modality) and questions regarding the patient (GAF score, psychiatric diagnoses). Therapists used the same code as the patient, which allowed matching the data. The code was encrypted before being saved to the data file.

Neither patients nor therapists received payment in return for their participation. The Cantonal Ethics Committee of Bern filed a letter of non-competence and stated no objection to the study (Req-2020-00421).

### Data Analysis

#### Quantitative Analyses

Data were screened for outliers, and three scores that were more than 3 standard deviations from the mean (one each on the following scales: CSQ, SEQ depth, and WAI-SR-T bond) were winsorized and replaced with the next lowest or highest value. All statistical analyses were conducted with IBM SPSS 26 (IBM^®^ SPSS^®^ Statistics Version 26). A significance level of .05 was set, and all *p* values are reported two-tailed. Because of the small sample size for the videoconferencing group, the results for this group are reported at a descriptive level only; for statistical analyses, the data from the videoconferencing group and the telephone group were collapsed into a telepsychiatry group. The main outcome variables were the SEQ, CSQ, WAI-SR-P, and WAI-SR-T. The sequential multiple regressions with *modality* as the predictor (dichotomized as face-to-face vs telepsychiatry) were adjusted for therapist and how many sessions the patient had already had in the current modality. According to post-hoc calculations with G*Power 3.1, achieved power to detect a medium effect with the current sample was good (.84). As for this study all patients who were eligible and attended a treatment session during the relevant time period were approached, a priori power calculations would not have been meaningful. For the analyses of patients’ preferences, chi square statistics and frequencies were calculated.

#### Qualitative Analyses

Patients’ and therapists’ responses to the open-ended questions were analysed for (1) factors that were perceived as positive (patient) or facilitated the session (therapist), and (2) factors that were perceived as negative (patient) or complicated the session (therapist). An inductive content analysis following the stages described by S Elo and H Kyngas [68] was conducted for each treatment modality group for responses from the patients and the therapists by one of the first authors. Content analysis allows large amounts of text to be organised into a smaller number of meaningful categories [69]. Topics referred to in several of the responses are reported in the result section.

## Results

### Preliminary Analyses

#### Reasons for Allocation to Treatment Modality

The following reasons were reported most frequently by the therapists for choosing a specific treatment modality: The modality was preferred by the patient (*n* = 39; similar frequency for face-to-face and telepsychiatry groups), the patient themselves or a member of their household belonged to the risk group (*n* = 9), and the patient needed to be seen in person because of the severity of the present condition (*n* = 4).

#### Sample Characteristics

Mean age of the overall sample was 39.10 (*SD* = 11.19) years, 55% were male, and they had attended an average of 2.42 (*SD* = 1.49) sessions in the current modality. The patients had 1.68 (*SD* = 0.65, range 1 to 3) psychiatric diagnoses on average (including the diagnosis of ADHD). The most common comorbidity was a F30-39 diagnosis (major mood disorders; 27%), followed by F40-49 (anxiety, stress-related, dissociative, and somatoform disorders; 22%), F10-19 (substance use related disorders; 7%), and F60-69 (personality disorders, impulse-control and “habit” disorders; 7%). Patients’ average GAF score was 68.62 (*SD* = 11.84). In terms of highest education, 5% had completed compulsory school, 27% had completed an apprenticeship, 25% had their school-leaving examination (a qualification to enter university), and 43% had a university degree. Details on sample characteristics by group can be seen from Table 1.

**Table 1.** Descriptive Information by Treatment Modality Group

|  | face-to-face<br>(*n* = 29) | videoconferencing<br>(*n* = 11) | telephone<br>(*n* = 20) | *p* (Cohen's *d*)<sup>a</sup> |
| --- | --- | --- | --- | --- |
| **Number of sessions** | 1.59 (0.87) | 2.91 (1.81) | 3.35 (1.39) | .000 (1.30)<sup>b</sup> |
| **Gender (% male)** | 48.3% | 63.6% | 60.0% | .311 (0.26)<sup>c</sup> |
| **Age** | 39.62 (11.22) | 38.00 (9.89) | 38.95 (12.26) | .730 (0.09)<sup>b</sup> |
| **Highest education** |  |  |  | .067 (–)<sup>d</sup> |
| Compulsory school | 3.4% | 9.1% | 5.0% |  |
| Apprenticeship | 27.6% | 27.3% | 25.0% |  |
| School-leaving examination | 24.1% | 18.2% | 30.0% |  |
| University degree | 44.8% | 45.5% | 40.0% |  |
| **No of psychiatric diagnoses (incl. ADHD)** | 1.66 (0.61) | 1.55 (0.52) | 1.80 (0.77) | .749 (0.08)<sup>b</sup> |
| **Comorbidities** |  |  |  |  |
| F10-19 | 6.9% | 0.0% | 10.0% | 1.000 (–)<sup>d</sup> |
| F30-39 | 34.5% | 18.2% | 20.0% | .185 (0.35)<sup>c</sup> |
| F40-49 | 20.7% | 36.4% | 15.0% | .859 (0.05)<sup>c</sup> |
| F60-69 | 0.0% | 0.0% | 20.0% | .113 (–)<sup>d</sup> |
| **GAF-score<sup>e</sup>** | 67.59 (11.92) | 72.55 (5.85) | 67.95 (14.02) | .519 (0.17)<sup>b</sup> |
| **COVID-19 high risk group** | 10.3% | 0.0% | 15.0% | 1.000 (–)<sup>d</sup> |
| **Impact of COVID-19** |  |  |  |  |
| change to everyday life | 3.62 (1.05) | 4.00 (1.33) | 4.10 (1.25) | .145 (0.38)<sup>b</sup> |
| distress | 2.83 (1.10) | 2.80 (1.32) | 3.25 (1.33) | .395 (0.22)<sup>b</sup> |
| fear | 2.38 (1.18) | 2.60 (0.84) | 2.70 (1.17) | .329 (0.25)<sup>b</sup> |

*Note:* The values represent the mean and the standard deviation are reported in brackets. School
leaving examinations are similar to A levels in the UK and qualify the individual to study at a
university. <sup>a</sup>To check for statistically significant a priori differences between groups, the face-to-face
group was compared to the collapsed telepsychiatry groups (telephone and videoconferencing)
because of the small sample size in the videoconferencing group. <sup>b</sup>Two-samples *t*-test. <sup>c</sup> $\chi^2$ test.
<sup>d</sup>Fisher's exact test. <sup>e</sup>As data was provided by two therapists, a second *t*-test with Z standardized GAF
scores (standardization within therapist) was conducted, which yielded similar results, $t(58) = -0.62$ ,
$p = .541$ , $d = 0.16$ .

#### A Priori Differences Between the Groups

Patients in the telepsychiatric group had attended statistically significantly more sessions in the present modality than patients in the face-to-face group, *t*(47.94) = −5.03, *p* < .001, *d* = 1.30. All other group differences, including those on three potential confounding variables considered, age, questions regarding patients’ experience with the COVID-19 pandemic, and GAF, did not reach statistical significance (see Table 1).

### Quantitative Analyses

#### SEQ, CSQ, WAI-SR-P, and WAI-SR-T

Table 2 presents an overview of the mean scores per group as well as the standardized regression weight of the *treatment modality* variable adjusted for *therapist* and *number of sessions* for each of the sequential multiple regressions (see Supplementary File 1, Tables B-K, for the full regression tables). The telepsychiatric sessions were rated as significantly less deep than the face-to-face sessions (*B* = −0.89, *p* = .001). While not statistically significant in step 1 (*B* = 0.01, *p* = .995), the predictor *number of sessions* approached significance after the predictor *modality* was introduced (*B* = 0.17, *p* = .062). We explored the association between number of sessions and SEQ depth in more detail. Individual scatterplots per group revealed a small, non-significant association between number of sessions and depth for the face-to-face group (*r* = .26, *p* = .168) and the telephone group (*r* = .10, *p* = .672). By contrast, patients in the videoconferencing group tended to experience more depth to their session the more sessions they had already had in that modality (*r* = .39, *p* = .243), an association that increased considerably after excluding an extreme value *(r* = .68, *p* = .030).

**Table 2.** Questionnaire Means and Standard Deviations per Treatment Modality Group and Adjusted Standardised Regression Coefficients

| | Face-to-face<br>(*n* = 29) | Video-<br>conferencing<br>(*n* = 11) | Telephone<br>(*n* = 20) | $\beta$ | *p* |
| --- | --- | --- | --- | --- | --- |
| **SEQ** |  |  |  |  |  |
| depth | 5.50 (0.73) | 4.47 (1.07) | 5.12 (0.89) | -0.49 | .001 |
| smoothness | 5.68 (1.02) | 5.14 (1.40) | 5.49 (1.05) | -0.10 | .508 |
| post-session positivity | 5.36 (1.12) | 5.35 (1.14) | 5.44 (0.98) | 0.06 | .731 |
| **CSQ** |  |  |  |  |  |
| total score | 3.75 (0.30) | 3.63 (0.38) | 3.71 (0.30) | -0.23 | .143 |
| **WAI-SR-P** |  |  |  |  |  |
| bond | 4.41 (0.51) | 4.39 (0.60) | 4.21 (0.72) | -0.16 | .322 |
| tasks | 3.99 (0.52) | 3.75 (0.68) | 3.78 (0.68) | -0.27 | .084 |
| goals | 3.95 (0.58) | 4.14 (0.49) | 3.93 (0.77) | 0.08 | .612 |
| **WAI-SR-T** |  |  |  |  |  |
| bond | 4.16 (0.50) | 4.30 (0.33) | 4.04 (0.53) | -0.19 | .207 |
| tasks | 3.75 (0.45) | 3.93 (0.37) | 3.61 (0.50) | -0.19 | .221 |
| goals | 3.62 (0.47) | 3.91 (0.44) | 3.53 (0.39) | -0.03 | .844 |

*Note:* The values represent the mean; the standard deviation is reported in brackets. The maximum scores for the SEQ subscales, the WAI-SR subscales, and the CSQ scale are 7, 5, and 4, respectively.
The standardized beta coefficient for the predictor *modality* (face-to-face or telepsychiatry (telephone and videoconferencing collapsed); reference group: face-to-face), which is adjusted for therapist and number of sessions, is reported for each outcome variable together with the *p* value of the regression coefficient.

None of the other models reached statistical significance, i.e. *modality* was not a statistically significant predictor for session smoothness, post-session positivity, satisfaction with the treatment received, or TA as rated by both patients and therapists.

### Patients’ self-reported preferences

Patients were asked whether they preferred the therapy sessions as they were before the COVID-19 pandemic, whether it made no difference to them, or whether they preferred the current sessions. Overall, 56.7% reported that they felt it made no difference, followed by 38.3% who preferred the sessions as they were before the pandemic. A chi-square analysis comparing the face-to-face group and the telepsychiatry group revealed no statistically significant differences in responses for the treatment modalities, *p* = .917 (Fisher’s Exact Test; see Table 3 for the figures for each of the groups).

**Table 3.**
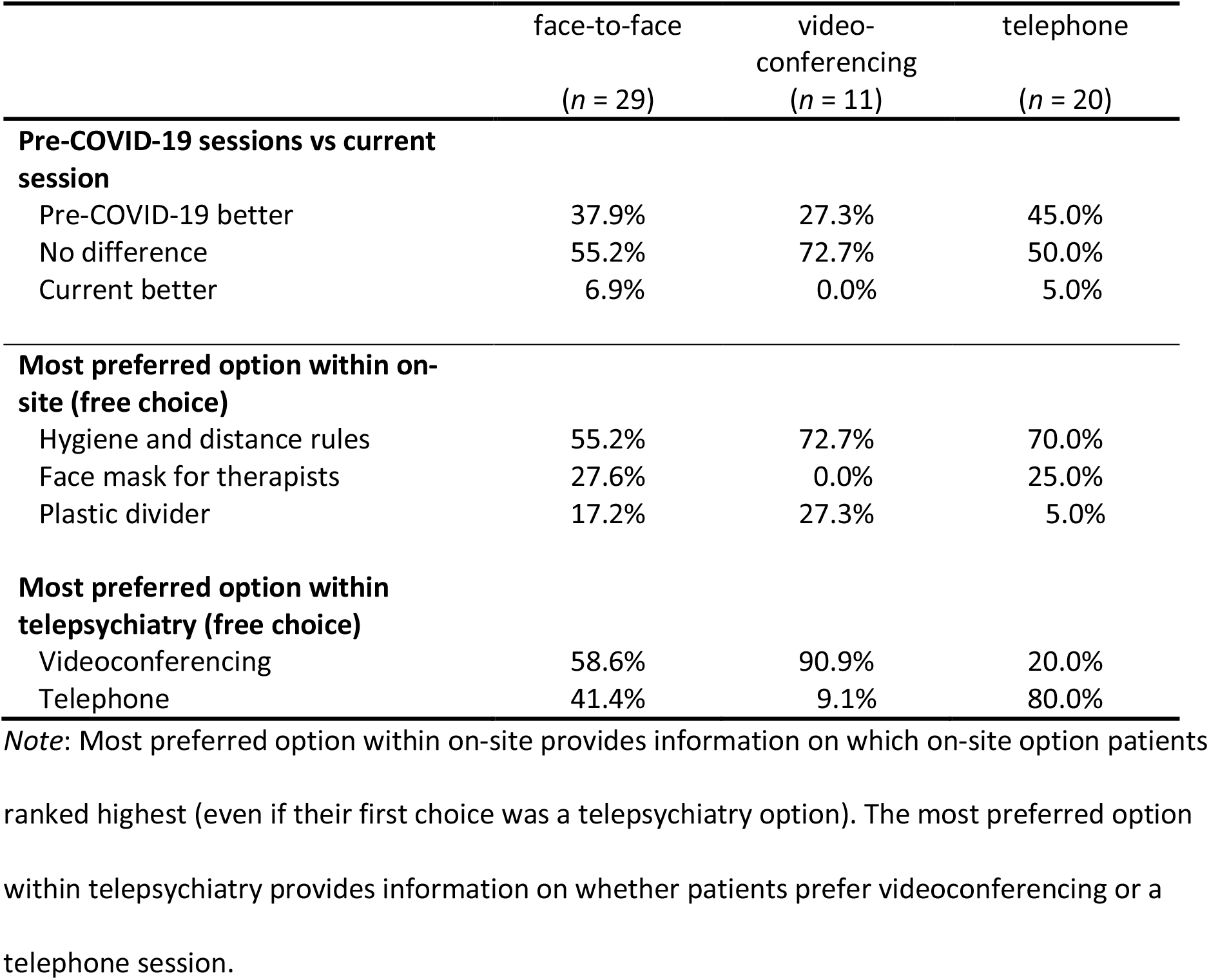
Preferences for pre-COVID-19 or Current Format and Patient Treatment Modality Preferences by Treatment Modality Group

|  | face-to-face<br>(n = 29) | video-<br>conferencing<br>(n = 11) | telephone<br>(n = 20) |
| --- | --- | --- | --- |
| **Pre-COVID-19 sessions vs current session** |  |  |  |
| Pre-COVID-19 better | 37.9% | 27.3% | 45.0% |
| No difference | 55.2% | 72.7% | 50.0% |
| Current better | 6.9% | 0.0% | 5.0% |
| **Most preferred option within on-site (free choice)** |  |  |  |
| Hygiene and distance rules | 55.2% | 72.7% | 70.0% |
| Face mask for therapists | 27.6% | 0.0% | 25.0% |
| Plastic divider | 17.2% | 27.3% | 5.0% |
| **Most preferred option within telepsychiatry (free choice)** |  |  |  |
| Videoconferencing | 58.6% | 90.9% | 20.0% |
| Telephone | 41.4% | 9.1% | 80.0% |

*Note:* Most preferred option within on-site provides information on which on-site option patients ranked highest (even if their first choice was a telepsychiatry option). The most preferred option within telepsychiatry provides information on whether patients prefer videoconferencing or a telephone session.

Participants were also asked about their treatment modality preferences (from most preferred to least preferred) if they were to have a choice among five options. The three groups differed in terms of their preferences, as can be seen from Figure 1. The face-to-face group had a strong preference for on-site treatment, although a majority would prefer sessions in compliance with hygiene and distance rules. In the telepsychiatry group, for about half of the patients their current modality was their most preferred one, whereas the other half would prefer on-site sessions, mostly in compliance with hygiene and distance rules. Analyses of the preferences within the face-to-face options revealed an overall preference for sessions following hygiene and distance rules only, whereas no clear preference could be identified in terms of videoconferencing vs telephone sessions (see Table 3).

**Figure 1.**
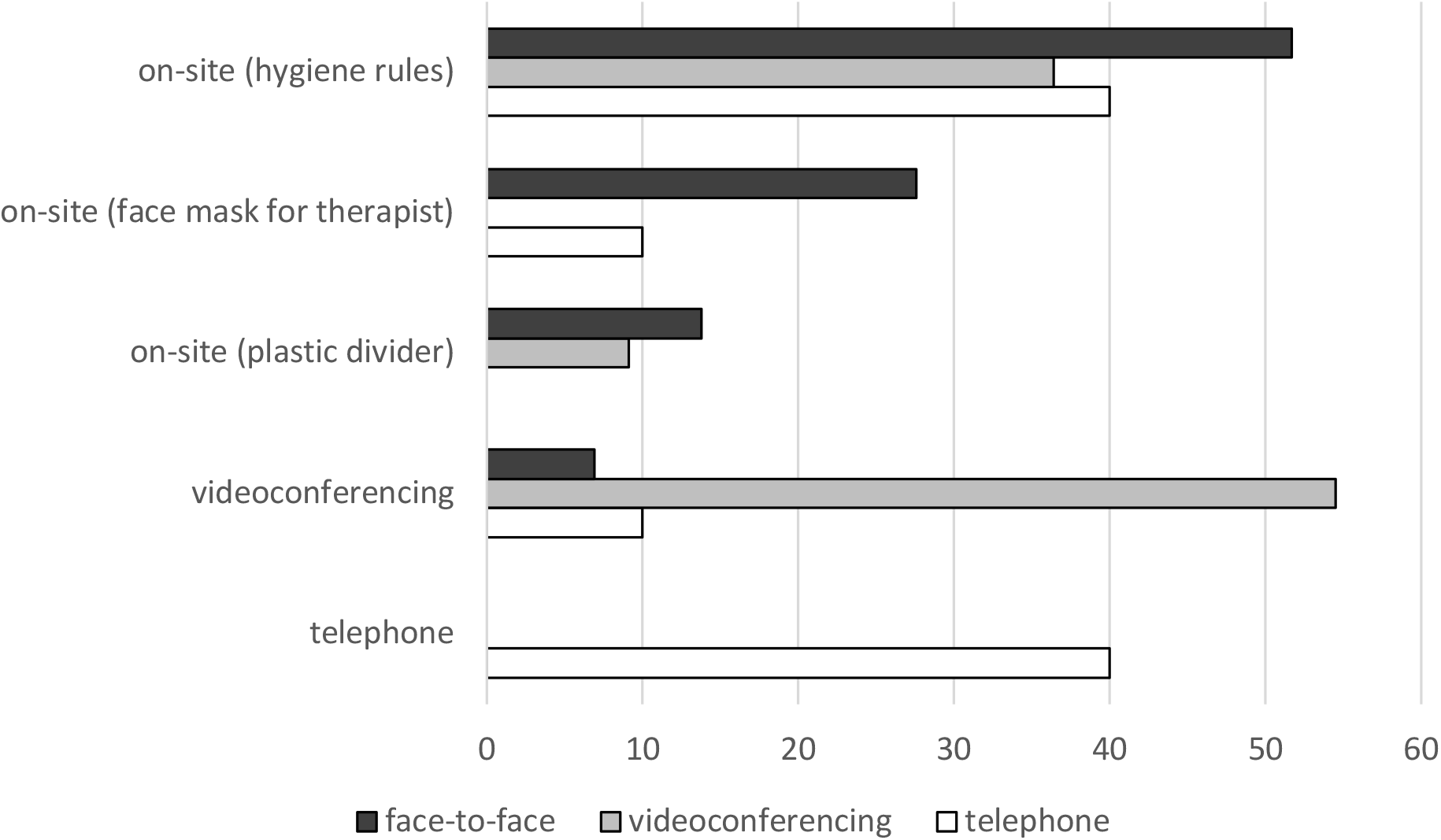
Preferred Modality If Patients Had Free Choice Among Five Options *Note*. This figure shows the percentage of patients in each group that indicated a specific option as their first choice.

### Qualitative Analyses

#### What did patients like and dislike about the specific session?

Although patients were asked what they liked and disliked about the specific session *modality*, the vast majority of responses were more general and seemed to refer to the session as a whole.

In the face-to-face group, most of the positive aspects mentioned concerned either the therapist’s behaviour (e.g. empathic, makes me feel taken seriously, friendly, understanding) or the therapy session itself (e.g. an individual aspect that patients found particularly useful about the session or being able to talk to the therapist in general). The most common issue identified with the specific session was that the mask hampered communication (e.g. no feedback from facial expressions, therapist harder to understand acoustically), which was mentioned by about one in five patients. The remaining points raised concerned administrative issues (e.g. having to wait, temporary relocation of therapy sessions to a different building).

Positive aspects identified by the videoconferencing group were related either to the therapist/therapy session itself or to organisational advantages of the modality (e.g. saving time, no waiting time if the therapist is running late). Some patients also mentioned that they appreciated having the opportunity to conduct a videoconferencing session, felt it was uncomplicated or rated it even on a par with face-to-face sessions. The following negative aspects were mentioned by two patients each: Technical problems (e.g. poor connection), lack of distance from everyday life (e.g. no journey to get to the session), and specific issues with the modality (e.g. more difficult to concentrate or feeling less present).

The majority of the telephone group’s positive comments again referred to the therapist or the therapy session. Some patients explicitly mentioned symptomatic and/or organisational advantages of this modality (e.g. being more focused on the session’s content because of the absence of visual cues, no need to use public transport, saving time). About one in five to six patients voiced concerns regarding the absence of visual input (e.g. no mimic feedback), felt that speaking over the phone was impersonal, or reported encountering administrative problems (e.g. poor sound quality). Circumstances identified as having the potential to make a telephone session more difficult were: not being sufficiently familiar with the therapist and the absence of mimic feedback when discussing specific (personal) topics. Conversely, patients felt that a telephone session worked well to discuss medication or when there was already an established bond of trust with their therapist.

#### Complicating and facilitating factors identified by therapists

Regarding face-to-face sessions, most of the complicating factors, which were mentioned for a little more than 1 out of 2 patients, were related to communication difficulties caused by the face mask (e.g. reduced access to facial expressions, acoustic communication problems, some patients feeling insecure or uncomfortable). For about two out of five patients, therapists felt that the sessions were fairly or fully comparable to the pre-COVID-19 sessions. A pre-existing bond of trust was reported to mitigate negative effects of wearing a mask, and therapists perceived the mask to be less of a problem for some patients than for others, although it was not possible to identify specific factors from the responses provided. Finally, in a small number of cases (1 out of 10 patients) therapists reported positive effects of wearing a face mask as some patients seemed more attentive during the session as a result.

The issues most commonly identified with videoconferencing sessions were: not being able to get a full picture of the patient (both figuratively and literally speaking; e.g. limited access to body language cues; about three out of four cases) and limitations in terms of therapeutic measures that could be employed (e.g. drawing up to-do lists, using whiteboards; raised for close to half of the cases). Only few technical issues were reported. A mitigating factor mentioned by the therapists was the main goal of the session; for instance, the limitations of a videoconferencing session were less important for sessions that focused on medication rather than psychotherapeutic work.

In terms of telephone sessions, the issues raised most frequently were the lack of access to body language and facial expression cues and the associated limited insight into patients’ affective or emotional reactions (about one out of three cases). In some cases, therapeutic measures were limited, or the therapist felt that patients seemed more distracted than in regular face-to-face sessions. In addition, telephone sessions were perceived as less suitable for patients who were very unwell and/or needed to discuss difficult topics. Conversely, speaking over the phone worked well for sessions that focused on medication. For about one in three cases, therapists reported that they felt the sessions were at least fairly comparable to pre-COVID-19 sessions and/or that the restrictions were not particularly problematic.

## Discussion

This study sought to explore adult ADHD patients’ and their therapists’ experience of a therapy session during the COVID-19 pandemic. Our results indicate that patients who attended a face-to-face session with the therapist wearing a face mask experienced significantly more depth to their session than patients who received telepsychiatric care. Descriptively, depth was lowest in the video-conferencing group, but further exploratory analyses suggested that depth in that group increased with the number of sessions. Thus, over time the difference in session depth between modalities might decline, which would be in line with the findings reported by Morgan and colleagues [70], who did not observe statistically significant differences in session depth for tele-mental health vs face-to-face psychological or psychiatric services. We can only speculate about possible explanations for an initially lower session depth rating in the telepsychiatry group. Patients might experience initial technical difficulties, need time to adapt to the new format, or need time to become accustomed to discussing personal or intimate topics during a telepsychiatric therapy session.

No statistically significant effects of treatment modality were observed for any of the other outcome variables. Post-session positivity did not differ between modalities, which suggests that there were no major differences in the effects of face-to-face and telepsychiatric sessions on patients’ post-session well-being. Also, patients’ satisfaction with the services received did not differ depending on modality, and no statistically significant differences in patients’ ratings of TA across the three conditions were observed. Although no direct comparison to pre-COVID-19 TA is possible, the results do not suggest that one type of adjustment made because of the pandemic might be particularly detrimental to TA as perceived by patients. The same seems to apply to TA as perceived by the therapists. Contrary to previous studies [see e.g. 33], no statistically significant difference in TA rating depending on treatment modality was observed. This difference to previous findings might be due to the fact that the therapists in the present study already knew their patients. According to SG Simpson and CL Reid [33], lower TA ratings for telepsychiatry were observed mainly in the early stages of an intervention. The findings from our study indicate that the modality chosen may have little bearing on TA for both patients and therapists. In sum, we observed few differences between face-to-face sessions (with the therapist wearing a face mask) and telepsychiatric sessions for adult patients with ADHD, and it seems possible that the difference that was observed would decrease as the number of sessions in the new modality increases.

In line with other findings reported thus far, two-thirds of all patients felt their sessions in the current treatment modality were not inferior to the pre-COVID-19 modality, and there was no statistically significant difference in that respect between patients in the face-to-face group and in the telepsychiatry group. If patients could choose freely from a number of options, however, over 60% of participants ranked on-site treatment with adherence to hygiene rules rather than on-site treatment with the therapist wearing a face mask or using a plastic divider as their first choice for on-site treatment. More than half of the patients in the face-to-face treatment group chose face-to-face treatment with adherence to hygiene and distance rules as their favourite option, as did the majority of patients in the telepsychiatry group whose first preference was an on-site option. Only a few of the patients in the face-to-face group opted for a telepsychiatric treatment modality as their first choice, whereas about half of the patients in the telepsychiatry group selected their current telepsychiatric treatment modality as their first choice. These findings reflect the fact that in many cases the choice of treatment modality was based on or in line with the patient’s preference.

Despite the fact that the majority of patients reported preferring a face-to-face modality that did not involve the therapist wearing a face mask, only about one in five patients in the face-to-face group actively mentioned issues related to the therapist wearing a mask (e.g. lack of facial expression cues or issues with acoustic comprehension) as something they disliked about their session. Rather, the results of the content analysis suggest that – across all conditions – patients’ experience of the session was mainly shaped by positive aspects related to the therapist, specific contents of the therapy session, or simply having the opportunity to speak to the therapist/attend a therapy session. The therapists, by contrast, mentioned issues relating to their wearing a mask in about half of the cases, although in a few cases they also reported some unexpected advantages of the face mask (e.g. patient appearing to be more focused or attentive). Thus, it seems that the absence of visual distractors (facial cues) might result in patients focusing more on acoustic input, i.e. the content of the session. It is possible that the therapists generally attached more importance to the question of the effects of the face mask than the patients did. Additionally, the open-ended questions for patients and therapists were formulated somewhat differently, which may have contributed to the therapists discussing the effects of wearing a mask more frequently. Either way, the fact remains that 80% of the patients in the face-to-face group did not feel the need to raise issues with the face mask.

In the telepsychiatric groups, in addition to the positive comments on the therapy session in general, patients additionally highlighted symptomatic and organisational advantages as something they liked about their session (e.g. no public transport, no journey, saving time). In the videoconferencing group, some experienced technical issues, emphasised a lack of distance from everyday life (e.g. because there was no journey to the session) or raised specific issues with the modality (e.g. feeling less present). In the telephone group, about one in five patients raised issues related to the lack of visual cues, and some felt the telephone session was somewhat impersonal or had encountered administrative issues. Similar to the face-to-face group, the therapists were again much more concerned with the limitations of the telepsychiatric modalities, particularly the full or partial lack of visual cues, than were their patients.

The responses to the open-ended questions on how the session was experienced also provided some insight into the circumstances under which a telepsychiatric session was considered more or less suitable. One aspect mentioned by both patients and therapists was session content. Telepsychiatric treatment was perceived to be more suitable if the session focused on discussing medication or other issues that were not too personal in nature. If more sensitive/intimate topics needed to be discussed, a face-to-face session was considered preferable. Therapists felt that telephone consultations were less appropriate for patients who were particularly unwell. Both patients and therapists also mentioned that, because of the limitations of telepsychiatry, some therapeutic methods could not be implemented (e.g. using white boards for to-do lists or plans), thereby highlighting another factor that may need to be considered when deciding on the modality of a session.

### Strengths, Limitations, and Future Research

Strengths of this study include that it sheds light on both the patients’ and the therapists’ experience with different treatment settings and provides us with qualitative information on these experiences. Moreover, to our knowledge it is the first study to investigate how an on-site session with the therapist wearing a face mask compares to telepsychiatric provision, both of which are potential adjustments that allow for continued treatment during a situation such as the COVID-19 pandemic. Yet, the generalisability of our results to other psychiatric disorders will need to be examined in future studies; depending on the predominant symptoms, patients may react differently to a scenario in which the therapist is wearing a face mask [see e.g. 11].

The focus on adult patients with ADHD is another strength of the study, as research on the suitability of telepsychiatric treatment for adult patients with ADHD is particularly scarce [51] and the various symptoms of ADHD, such as inattentiveness and organisational deficits, have distinct potential to interfere with this type of treatment modality. Due to ADHD’s high prevalence in adults, often demanding continuous treatment, and its frequent presentation with severe psychiatric co-morbidities, patients with ADHD are a particularly important group to study. This research offers a valuable contribution to that still understudied topic. While our ADHD sample was comparable in terms of certain aspects such as comorbidity of major mood-disorders [e.g. 71], the comorbidity of substance abuse was lower than what has been reported in other studies [72, 73]. Moreover, the number of patients with high educational achievements was above average. These aspects should be taken into account in terms of the generalisability of the findings to other patients with ADHD.

Randomized allocation to treatment modality was not possible in this study. We identified a number of potential confounding variables and examined whether there were any group differences for these variables. Patient age was included because younger patients may be more familiar and confident with the use of telecommunication, which could have distorted the results in case of systematic differences between the three groups. The self-reported extent to which the patient was affected by the COVID-19 pandemic could be related to treatment modality (e.g. patients who were more affected were also more likely to stay at home and receive tele-psychiatric care) and at the same time be associated with the outcome variables (e.g. patients who are more affected by the pandemic may feel more unwell, which may affect how they experience their therapy sessions). Finally, level of functioning could also be related to both treatment modality and our outcome variables. However, for none of these variables did we observe statistically significant differences between groups. Still, the findings of the present study are limited to situations in which patients (1) are already under treatment and familiar with their therapist, and (2) have a say in the choice of adjustment that is made in order to continue treatment during a situation such as the COVID-19 pandemic. It is likely that many patients chose a setting they thought they would feel comfortable with and/or would suit them for one reason or another. Although this does not rule out that patients still face difficulties with the modality chosen (e.g. problems with focusing during a telepsychiatric session), it is conceivable that patients who do not have a choice in the first place would find it comparably more difficult to adapt to their new setting, a question that should be addressed in future research. Nonetheless, the circumstances in the study at hand reflect a real-life situation and our study provides valuable insights into how certain treatment options compare and what may need to be considered when deciding on which adjustments should be made.

The fact that all patients were treated by one of two therapists also limits the generalisability of the results. Future research using a broader sample of therapists will be needed to check the replicability of the results. Moreover, while the present study provides insight into the vital question of whether interim telepsychiatric treatment might affect TA very differently from a face-to-face session with the therapist wearing a face mask, future studies should establish a baseline TA (i.e. TA during therapy sessions without any adjustments) to investigate how individual adjustments affect TA. In addition, the impact of a prolonged use of certain adjustments or modalities on patient and therapist experience remains unclear. Finally, because of the small number of cases in the video-conferencing group, the groups were collapsed for the main analyses. While descriptive data in many cases do not suggest major differences between the telephone and the video-conferencing group for most variables, future studies should aim to include large samples to increase power to detect differences not only between a face-to-face and a telepsychiatry group but also between different types of telepsychiatric settings.

### Implications for Practice and Conclusion

Continuing psychiatric or psychotherapeutic treatment during the COVID-19 pandemic required significant changes to the way therapy sessions were conducted. First findings on the impact of such adjustments reported in the study at hand are encouraging; both options – on-site with the therapist wearing a mask and telepsychiatry – seem viable alternatives for continuing therapies during a situation such as the COVID-19 pandemic. Although telepsychiatric sessions, particularly those involving videoconferencing, were experienced as less deep by the participants, no differences were observed in post-session positivity, and the difference concerning depth might decline over time. Also, no differences in terms of client satisfaction or TA as perceived by both patient and therapist were observed. While the majority of patients would prefer an on-site solution that does not involve the therapist wearing a face mask, patients’ comments on the on-site session focused more on positive elements such as being able to attend the session/speak to their therapist rather than on issues related to the therapist wearing a face mask. Therapists seemed to be more concerned about potential negative impacts of the masks, and a similar observation was made regarding the lack of visual cues in telephone sessions.

When deciding on the modality or adjustment for a specific session, our findings suggest that it may be worthwhile for clinicians to consider the following two questions in addition to feasibility and patient preference: First, is the topic of the session highly personal in nature? If so, it might be worth discussing with the patient whether they would feel more comfortable with a face-to-face session. Conversely, if the topic is more general in nature (e.g. discussing medication), a telepsychiatric solution could be considered. Second, does the session contain therapeutic elements, such as drawing up plans and using whiteboards, that may be difficult or impossible to implement in a telepsychiatric setting and would not having access to these elements be particularly detrimental to the session (aims)? This is particularly relevant to sessions over the phone; advances in technology may make this question obsolete if videoconferencing is used and both parties are technically sufficiently skilled. Finally, some patients mentioned a lack of (physical and/or temporal) distance to their everyday life if the session was off-site. It may therefore be helpful to advise patients to take a short break (e.g. from working from home) prior to their telepsychiatric appointment or go to a specific location (e.g. separate room or place) when participating in a therapy session.

## Data Availability

The datasets generated and analysed during the current study are available from the corresponding author on reasonable request.

## Declarations

### Ethics approval and consent to participate

All participants provided electronic informed consent. The research was carried out in accordance with the Declaration of Helsinki. The study was submitted to the Cantonal Ethics Committee of Bern (Req-2020-00421) under the Federal Act on Research involving Human Beings. The Committee concluded that there were no concerns with the study with regards to the Human Research Act and filed a letter of non-competence. This declaration of non-competence corresponds to a declaration of no-objection under Swiss legislation regulating research involving human beings [see also e.g. 1, 2].

### Consent for publication

Not applicable.

### Availability of data and materials

The datasets generated and analysed during the current study are available from the corresponding author on reasonable request in order to protect anonymity of the participants (in-depth responses to open-ended questions).

### Authors’ contributions

HW/ML/AB/PB: conception of the study; HW/ML: ethics; HW: preparation of online questionnaires; AB/PB: participant recruitment; HW/VA: data analysis; HW/ML: writing of the first draft of the manuscript; AB/ES/PB/SY/VA: writing and revisions. All authors read and approved the final version of the manuscript.

## Acknowledgments

We would like to thank Heather Murray for copy-editing the manuscript.

## List of Abbreviations

ADHD: attention-deficit/hyperactivity disorder
CSQ: Client Satisfaction Questionnaire
GAF: Global Assessment of Functioning
SEQ: Session Evaluation Questionnaire
TA: therapeutic alliance
WAI-SR-P: Working Alliance Inventory short revised therapist version
WAI-SR-T: Working Alliance Inventory short revised patient version

## Footnotes

1 This means most of the data collection took place during the extraordinary situation, which lasted from the 16^th^ of March until the 19^th^ of June.

2 Due to a technical error, item 6 was missing from the questionnaire, which means that the subscale *smoothness* consisted of 4 instead of 5 items.

