## Supplementary File 1 for "Treatment provision for adults with ADHD during the COVID-19 pandemic: An exploratory study on patient and therapist experience with on-site sessions using face masks vs. telepsychiatric sessions"

**Table A**

*Overview of self-developed questions used in the study (German original wording and English translation)*

| Item | German original | English translation |
|---|---|---|
| C | Anleitung: Bitte beantworten Sie die vier folgenden Fragen dazu, wie Sie die COVID-19 Pandemie erleben.<br><br>Antwortoptionen C1-C4:<br>- *überhaupt nicht (1)*<br>- *(2)*<br>- *(3)*<br>- *(4)*<br>- *sehr stark (5)* | Instruction: Please answer the following four questions about how you are experiencing the COVID-19 pandemic.<br><br>Response options C1-C4:<br>- *not at all (1)*<br>- *(2)*<br>- *(3)*<br>- *(4)*<br>- *very much (5)* |
| C1 | Gehören Sie zu jenen Personen, die ein erhöhtes Risiko für einen schwereren Verlauf einer COVID-19-Erkrankung haben (dies sind Personen ab 65 Jahren oder Personen mit mindestens einer der folgenden Vorerkrankungen: Bluthochdruck, chronische Atemwegserkrankungen, Diabetes, Erkrankungen und Therapien, die das Immunsystem schwächen, Herz-Kreislauf-Erkrankungen, Krebs)? | Do you belong to the group of people who are at a higher risk of developing more severe illness from COVID-19 (these are people aged 65 and over or people with at least one of the following pre-existing conditions: High blood pressure, chronic respiratory disease, diabetes, diseases and therapies that weaken the immune system, cardiovascular disease, cancer)? |
| C2 | In welchem Ausmass hat die COVID-19 Pandemie Ihren Alltag verändert? | To what extent has the COVID-19 pandemic affected your everyday life? |
| C3 | Inwiefern fühlen Sie sich wegen der COVID-19 Pandemie gestresst? | To what extent do you feel distressed because of the COVID-19 pandemic? |
| C4 | Inwiefern löst die COVID-19 Pandemie bei Ihnen Angst aus? | To what extent does the COVID-19 pandemic trigger fear? |
| OP1 | Gab es etwas an der heutigen Sitzungsform, das Ihnen besonders gefallen hat?<br>*Offene Frage* | Was there anything you particularly liked about the way today's session took place?<br>*Open-ended question* |
| OP2 | Gab es etwas an der heutigen Sitzungsform, das Sie gestört hat?<br>*Offene Frage* | Was there anything you disliked about or felt uncomfortable with in the way today's session took place?<br>*Open-ended question* |
| OT3 | Wie bewerten Sie die aktuell durchgeführte Therapieform im Vergleich zu der normalerweise durchgeführten Therapieform bei diesem Patienten/dieser Patientin?<br>*Offene Frage* | For this patient, how do you evaluate the way the therapy sessions are currently conducted compared to the way they normally take place?<br>*Open-ended question* |
| F1 | Wie gefällt Ihnen die Art und Weise, wie die heutige Sitzung stattgefunden hat, im Vergleich zu den «normalen» | How do you like the way today's session took place compared to the "normal" therapy sessions before the pandemic outbreak (in |

| | | |
|---|---|---|
| | Therapiesitzungen vor dem Pandemieausbruch (persönlich vor Ort und ohne Schutzmassnahmen)?<br>- *Die «normalen» Therapiesitzungen waren besser*<br>- *Die Sitzungen so wie heute sind besser*<br>- *Für mich macht es keinen Unterschied* | person, on site and without protective measures)?<br>- *The "normal" therapy sessions were better*<br>- *The sessions as the one today are better*<br>- *It makes no difference to me* |
| F2 | Bitte ordnen Sie die folgenden Sitzungsmöglichkeiten so an, dass ganz oben jene Möglichkeit ist, die Sie sich am meisten wünschen würden (=1) und ganz unten jene Möglichkeit ist, die Sie sich am wenigsten wünschen würden (=5).<br>- *Telefon*<br>- *Videotelefonie (z.B. Jitsi oder Skype)*<br>- *Persönlich vor Ort, als Schutzmassnahme werden die gängigen Hygieneregeln beachtet (kein Händeschütteln, mindestens 2 Meter Abstand)*<br>- *Persönlich vor Ort, Therapeut/in trägt als Schutzmassnahme eine Schutzmaske*<br>- *Persönlich vor Ort, als Schutzmassnahme hat es eine Trennscheibe zwischen mir und dem/der Therapeuten/Therapeutin* | Please arrange the following options in a way that the most preferred is on top (=1) and the least preferred is at the bottom (=5).<br>- *Telephone*<br>- *Videoconferencing (e.g. Jitsi or Skype)*<br>- *In person and on site, as protective measures the usual hygiene rules are observed (no handshaking, at least 2 meters distance)*<br>- *In person and on site, therapist wears a face mask as a protective measure*<br>- *In person and on site, as a protective measure a plastic divider is situated between me and the therapist* |

*Note*: The SEQ, CSQ, WAI, and GAF, which were also used in this study, are pre-existing questionnaires. The relevant references can be found in the main manuscript.

**Table B**

*Hierarchical Regression Results for SEQ Depth*

| Variable | *B* | 95% CI for *B* | | *SE B* | β | $R^2$ | $\Delta R^2$ |
|---|---|---|---|---|---|---|---|
| | | *LL* | *UL* | | | | |
| Step 1 | | | | | | .01 | .00 |
| Constant | 5.25*** | 4.70 | 5.80 | 0.28 | | | |
| Therapist | -0.18 | -0.68 | 0.32 | 0.25 | -.10 | | |
| No of sessions | 0.01 | -0.16 | 0.18 | 0.09 | .02 | | |
| Step 2 | | | | | | .18* | .13 |
| Constant | 5.25*** | 4.72 | 5.77 | 0.26 | | | |
| Therapist | -0.10 | -0.57 | 0.36 | 0.23 | -.06 | | |
| No of sessions | 0.18 | -0.01 | 0.37 | 0.09 | .28 | | |
| Modality | -0.87** | -1.42 | -0.32 | 0.28 | -.46 | | |

*Note*: Modality: 0 = face-to-face, 1 = telepsychiatry. *$p < .05$. **$p < .01$. ***$p < .001$.

**Table C**

*Hierarchical Regression Results for SEQ Smoothness*

| Variable | *B* | 95% CI for *B* | | *SE B* | β | $R^2$ | $\Delta R^2$ |
|---|---|---|---|---|---|---|---|
| | | *LL* | *UL* | | | | |
| Step 1 | | | | | | .09 | .06 |
| Constant | 6.08*** | 5.44 | 6.71 | 0.32 | | | |
| Therapist | -0.65* | -1.21 | -0.09 | 0.28 | -.30 | | |
| No of sessions | -0.10 | -0.29 | 0.09 | 0.10 | -.14 | | |
| Step 2 | | | | | | .10 | .05 |
| Constant | 6.08*** | 5.44 | 6.71 | 0.32 | | | |
| Therapist | -0.63* | -1.19 | -0.06 | 0.28 | -.29 | | |
| No of sessions | -0.06 | -0.29 | 0.17 | 0.11 | -.08 | | |
| Modality | -0.22 | -0.89 | 0.45 | 0.33 | -.10 | | |

*Note*: Modality: 0 = face-to-face, 1 = telepsychiatry. $^{*}p < .05$. $^{**}p < .01$. $^{***}p < .001$.

**Table D**

*Hierarchical Regression Results for SEQ Positivity*

| Variable | *B* | 95% CI for *B* | | *SE B* | β | $R^2$ | $\Delta R^2$ |
|---|---|---|---|---|---|---|---|
| | | *LL* | *UL* | | | | |
| Step 1 | | | | | | .00 | .00 |
| Constant | 5.49*** | 4.86 | 6.13 | 0.32 | | | |
| Therapist | -0.12 | -0.69 | 0.44 | 0.28 | -.06 | | |
| No of sessions | -0.02 | -0.21 | 0.17 | 0.10 | -.03 | | |
| Step 2 | | | | | | .01 | .00 |
| Constant | 5.49*** | 4.85 | 6.13 | 0.32 | | | |
| Therapist | -0.14 | -0.71 | 0.44 | 0.29 | -.06 | | |
| No of sessions | -0.04 | -0.27 | 0.19 | 0.12 | -.06 | | |
| Modality | 0.12 | -0.56 | 0.79 | 0.34 | -.06 | | |

*Note*: Modality: 0 = face-to-face, 1 = telepsychiatry. $^{*}p < .05$. $^{**}p < .01$. $^{***}p < .001$.

**Table E**

*Hierarchical Regression Results for CSQ*

| Variable | *B* | 95% CI for *B* | | *SE B* | β | $R^2$ | $\Delta R^2$ |
|---|---|---|---|---|---|---|---|
| | | *LL* | *UL* | | | | |
| Step 1 | | | | | | .01 | .00 |
| Constant | 3.70*** | 3.51 | 3.88 | 0.09 | | | |
| Therapist | -0.03 | -0.20 | 0.13 | 0.08 | -.05 | | |
| No of sessions | 0.01 | -0.04 | 0.07 | 0.03 | .07 | | |
| Step 2 | | | | | | .05 | .00 |
| Constant | 3.79*** | 3.51 | 3.88 | 0.09 | | | |
| Therapist | -0.02 | -0.19 | 0.15 | 0.08 | -.03 | | |
| No of sessions | 0.04 | -0.03 | 0.11 | 0.03 | .20 | | |
| Modality | -0.14 | -0.34 | 0.05 | 0.10 | -.23 | | |

*Note*: Modality: 0 = face-to-face, 1 = telepsychiatry. $^{*}p < .05$. $^{**}p < .01$. $^{***}p < .001$.

**Table F**

*Hierarchical Regression Results for WAI Bond (Patient)*

| Variable | *B* | 95% CI for *B* | | *SE B* | β | $R^2$ | $\Delta R^2$ |
|---|---|---|---|---|---|---|---|
| | | *LL* | *UL* | | | | |
| Step 1 | | | | | | .00 | .00 |
| Constant | 4.33*** | 3.97 | 4.69 | 0.18 | | | |
| Therapist | 0.02 | -0.30 | 0.34 | 0.16 | .01 | | |
| No of sessions | 0.00 | -0.11 | 0.11 | 0.05 | .00 | | |
| Step 2 | | | | | | .02 | .00 |
| Constant | 4.34*** | 3.97 | 4.70 | 0.18 | | | |
| Therapist | 0.03 | -0.29 | 0.36 | 0.16 | .03 | | |
| No of sessions | 0.03 | -0.10 | 0.16 | 0.07 | .09 | | |
| Modality | -0.19 | -0.57 | 0.19 | 0.19 | -.16 | | |

*Note*: Modality: 0 = face-to-face, 1 = telepsychiatry. $^{*}p < .05$. $^{**}p < .01$. $^{***}p < .001$.

**Table G**

*Hierarchical Regression Results for WAI Tasks (Patient)*

| Variable | *B* | 95% CI for *B* | | *SE B* | β | $R^2$ | $\Delta R^2$ |
|---|---|---|---|---|---|---|---|
| | | *LL* | *UL* | | | | |
| Step 1 | | | | | | .01 | .00 |
| Constant | 3.93*** | 3.57 | 4.29 | 0.18 | | | |
| Therapist | -0.12 | -0.45 | 0.20 | 0.16 | -.10 | | |
| No of sessions | 0.00 | -0.11 | 0.11 | 0.05 | .01 | | |
| Step 2 | | | | | | .06 | .01 |
| Constant | 3.94*** | 3.58 | 4.29 | 0.18 | | | |
| Therapist | -0.10 | -0.41 | 0.22 | 0.16 | -.08 | | |
| No of sessions | 0.06 | -0.06 | 0.19 | 0.06 | .16 | | |
| Modality | -0.33 | -0.70 | 0.05 | 0.19 | -.27 | | |

*Note*: Modality: 0 = face-to-face, 1 = telepsychiatry. $^{*}p < .05$. $^{**}p < .01$. $^{***}p < .001$.

**Table H**

*Hierarchical Regression Results for WAI Goals (Patient)*

| Variable | *B* | 95% CI for *B* | | *SE B* | β | $R^2$ | $\Delta R^2$ |
|---|---|---|---|---|---|---|---|
| | | *LL* | *UL* | | | | |
| Step 1 | | | | | | .00 | .00 |
| Constant | 3.99*** | 3.61 | 4.37 | 0.19 | | | |
| Therapist | 0.05 | -0.29 | 0.38 | 0.17 | .04 | | |
| No of sessions | -0.02 | -0.13 | 0.10 | 0.06 | -.04 | | |
| Step 2 | | | | | | .01 | .00 |
| Constant | 3.99*** | 3.61 | 4.37 | 0.19 | | | |
| Therapist | 0.04 | -0.30 | 0.38 | 0.17 | .03 | | |
| No of sessions | -0.04 | -0.17 | 0.10 | 0.07 | -.08 | | |
| Modality | 0.10 | -0.30 | 0.50 | 0.20 | .08 | | |

*Note*: Modality: 0 = face-to-face, 1 = telepsychiatry. $^{*}p < .05$. $^{**}p < .01$. $^{***}p < .001$.

**Table I**

*Hierarchical Regression Results for WAI Bond Therapist*

| Variable | *B* | 95% CI for *B* | | *SE B* | β | $R^2$ | $\Delta R^2$ |
|---|---|---|---|---|---|---|---|
| | | *LL* | *UL* | | | | |
| Step 1 | | | | | | $.11^{*}$ | .08 |
| Constant | $3.85^{***}$ | 3.57 | 4.13 | 0.14 | | | |
| Therapist | $0.29^{*}$ | 0.04 | 0.54 | 0.12 | .30 | | |
| No of sessions | 0.06 | -0.02 | 0.15 | 0.04 | .19 | | |
| Step 2 | | | | | | $.13^{*}$ | .09 |
| Constant | $3.85^{***}$ | 3.58 | 4.13 | 0.14 | | | |
| Therapist | $0.31^{*}$ | 0.06 | 0.55 | 0.12 | .32 | | |
| No of sessions | 0.10 | 0.00 | 0.20 | 0.05 | .30 | | |
| Modality | -0.18 | -0.47 | 0.11 | 0.14 | -.19 | | |

*Note*: Modality: 0 = face-to-face, 1 = telepsychiatry. $^{*}p < .05$. $^{**}p < .01$. $^{***}p < .001$. *F* change was not significant for Step 2, which means that the inclusion of condition does not statistically significantly improve prediction of the outcome variable. As the outcome is based on the ratings from two therapists, the regressions were also run for each therapist individually, in which case *therapist* was removed as a predictor in Step 1. *Modality* was not a statistically significant predictor in either of the two hierarchical regressions.

**Table J**

*Hierarchical Regression Results for WAI Tasks Therapist*

| Variable | *B* | 95% CI for *B* | | *SE B* | β | $R^2$ | $\Delta R^2$ |
|---|---|---|---|---|---|---|---|
| | | *LL* | *UL* | | | | |
| Step 1 | | | | | | .10 | .06 |
| Constant | 3.48*** | 3.21 | 3.74 | 0.13 | | | |
| Therapist | 0.25* | 0.02 | 0.49 | 0.12 | .28 | | |
| No of sessions | 0.06 | -0.02 | 0.14 | 0.04 | .19 | | |
| Step 2 | | | | | | .12 | .07 |
| Constant | 3.48*** | 3.22 | 3.74 | 0.13 | | | |
| Therapist | 0.27* | 0.03 | 0.50 | 0.12 | .29 | | |
| No of sessions | 0.09 | 0.00 | 0.18 | 0.05 | .29 | | |
| Modality | -0.17 | -0.44 | 0.11 | 0.14 | -.19 | | |

*Note*: Modality: 0 = face-to-face, 1 = telepsychiatry. $^{*}p < .05$. $^{**}p < .01$. $^{***}p < .001$. As the outcome is based on the ratings from two therapists, the regressions were also run for each therapist individually, in which case *therapist* was removed as a predictor in Step 1. *Modality* was not a statistically significant predictor in either of the two hierarchical regressions.

**Table K**

*Hierarchical Regression Results for WAI Goals Therapist*

| Variable | *B* | 95% CI for *B* | | *SE B* | β | $R^2$ | $\Delta R^2$ |
|---|---|---|---|---|---|---|---|
| | | *LL* | *UL* | | | | |
| Step 1 | | | | | | .05 | .02 |
| Constant | 3.46*** | 3.20 | 3.72 | 0.13 | | | |
| Therapist | 0.19 | -0.04 | 0.43 | 0.12 | .22 | | |
| No of sessions | 0.04 | -0.04 | 0.12 | 0.04 | .12 | | |
| Step 2 | | | | | | .05 | .00 |
| Constant | 3.46*** | 3.19 | 3.73 | 0.13 | | | |
| Therapist | 0.20 | -0.04 | 0.43 | 0.12 | .22 | | |
| No of sessions | 0.04 | -0.05 | 0.14 | 0.05 | .14 | | |
| Modality | -0.03 | -0.31 | 0.25 | 0.14 | -.03 | | |

*Note*: Modality: 0 = face-to-face, 1 = telepsychiatry. *$p < .05$. **$p < .01$. ***$p < .001$. As the outcome is based on the ratings from two therapists, the regressions were also run for each therapist individually, in which case *therapist* was removed as a predictor in Step 1. *Modality* was not a statistically significant predictor in either of the two hierarchical regressions.
